# The Outcome of Gynecologic Cancer Patients With Covid-19 Infection: A Systematic Review And Meta-Analysis

**DOI:** 10.1101/2022.03.20.22272676

**Authors:** I Gde Sastra Winata, Januar Simatupang, Arie A Polim, Yakob Togar, Advenny Elisabeth Tondang

**Author notes:** **Correspondence to: Yakob Togar, MD.** Jl.Haji Achyar No 10. Duren Sawit, Jakarta 13440, Indonesia. **Disclaimers:** None Declared.

## Abstract

**Objective:** Cancer is a comorbidity that leads to progressive worsening of Covid-19 with increased mortality. This is a systematic review and meta-analysis to yield evidence of adverse outcomes of Covid-19 in gynecologic cancer.

**Methods:** Searches through PubMed, Google Scholar, ScienceDirect, and medRxiv to find articles on the outcome of gynecologic cancer with Covid-19 (24 July 2021-19 February 2022). Newcastle-Ottawa Scale tool is used to evaluate the quality of included studies. Pooled odds ratio (OR), 95% confidence interval (CI), random-effects model were presented. This study was registered to PROSPERO (CRD42021256557).

**Results:** We accepted 51 studies (1991 gynecologic cancer with Covid-19). Covid-19 infection was lower in gynecologic cancer vs hematologic cancer (OR 0.71, CI 0.56-0.90, *p* 0.005). Severe Covid and death were lower in gynecologic cancer vs lung and hematologic cancer (OR 0.36, CI 0.16-0.80, *p* 0.01), (OR 0.52, CI 0.44-0.62, *p* <0.0001), (OR 0.26, CI 0.10-0.67 *p* 0.005), (OR 0.63, CI 0.47-0.83, *p* 0.001) respectively. Increased Covid death is seen in gynecologic cancer vs breast, non-covid cancer, and non-cancer covid (OR 1.50, CI 1.20-1.88, *p* 0.0004), (OR 11.83, CI 8.20-17.07, *p* <0.0001), (OR 2.98, CI 2.23-3.98, *p* <0.0001) respectively.

**Conclusion:** Gynecologic cancer has higher Covid-19 adverse outcomes compared to non-cancer, breast cancer, non-metastatic, and Covid-19 negative population. Gynecologic cancer has fewer Covid-19 adverse outcomes compared to other cancer types, lung cancer, and hematologic cancer. These findings may aid health policies and services during the ongoing global pandemic.

## INTRODUCTION

The Covid-19 pandemic has changed the way of health care providers around the world manage care provided to their patients. The pandemic has also proven to shift the attitude of standard practice and procedure between providers and patients, for example, to reduce gynecologic cancer patients visiting the hospital as possible because the risk of getting infected with Covid-19 is increased regarding their comorbidities.^1^ Despite this circumstance, gynecologic cancer patients is still often required to perform routine hospital visits for treatments or other medical procedures under guidance made by gynecological cancer societies during the Covid-19 pandemic.^2^ The cancer incidence and mortality are still increasing around the world. According to Global Cancer Statistic: 2020 for gynecologic cancer, there are 604.127, 417.367, 313.959, 45.240, and 17.908 new cases of cancer of the cervix uteri, corpus uteri, ovary, vulva, and vagina respectively.^3^ Most concerns are coming from these patients about how they may proceed to seek or continue their cancer treatment and surveillance during the Covid-19 pandemic time whether they should continue or delay.^4^ Studies are showing various results on increased mortality and severity among cancer patients infected with Covid-19. Systematic review and meta-analysis studying the outcome of cancer patients with Covid-19 show 2.1-4% proportion of cancer patients among those infected with Covid-19, additionally compared to non-cancer with Covid-19 greater amount of mortality and severity are observed in cancer population with Covid-19.^5-7^ However studies and data on the outcome of gynecologic cancer patients with Covid-19 are still lacking. We are now entering the third year of the Covid-19 pandemic after the first confirmed case was announced in December 2019 in Wuhan, Hubei Province, The People’s Republic of China. Several SARS-CoV-2 variants of concerns listed by WHO (World Health Organization) pose challenges in mitigating the pandemic as these variants often increase transmission rate and severity.^8^ The world has been experiencing a wave of active case surges by these variants and on 26 November 2021 the WHO designated the variant Omicron (B.1.1.529) as an addition to the list.^9^ Thus we attempt to review the literature and quantify the effect of the SARS-Cov-2/Covid-19 infection among gynecologic cancer patients whether the risk of infection, hospitalization, severity, and mortality are increased than non-gynecologic cancer population.

## MATERIALS AND METHODS

We conducted this systematic review and meta-analysis according to the Preferred Reporting Items for Systematic Reviews and Meta-Analyses/PRISMA statement.^10^ This study and its protocol were registered to PROSPERO (CRD42021256557).

### Eligibility Criteria

We took into consideration of studies with observational cohort studies, case-control, cross-sectional, case report, and case series designs that evaluate the outcome of gynecologic cancer patients infected with Covid-19 from the year 2019. Each study ought to report Covid-19 associated infection, hospital admission, mortality, severity, or admission to the ICU provided both in the main result or supplementary data. We exclude studies other than the English language, reviews or guidelines, and inconceivable results of the sought outcome.

### Comparator(s) / Control

Non-cancer Covid-19 patients, non-Covid-19 cancer patients, other cancer types / non-gynecological cancer with Covid-19.

### Database and Literature Search

Study articles were systematically searched through PubMed/Medline, ScienceDirect, Google Scholar, and medRxiv. Relevant articles had been screened from 24 July 2021 to 19 February 2022. Reference searches from retrieved articles citation lists were identified if any were needed. Boolean operators technique used for Pubmed/Medline search with (“COVID-19” or “2019-nCoV” or “SARS-CoV” or SARSCOV2 or 2019-nCov or “2019 coronavirus” or covid19) AND (gynecology or gynaecology) AND (tumor or malignancy or cancer) AND (outcomes or outcome) AND (gyn* tum* or gyn *malign* or gyn* cancer) AND (cancer surgery or oncolog* surger*) AND (brachytherapy or radiotherapy). We used “Gynecologic cancer AND Covid-19” with Google Scholar, Science Direct, and medRxiv. Two authors separately handled the literature search. Findings were accumulated and stored in Mendeley and Zotero for management and automated duplicate identification. Thorough stepwise screening from title and abstract was then conducted to determine possible article inclusion. Potentially eligible studies were then evaluated for in-depth full-text review. Each author would consult senior authors to resolve any differences found during the literature’s selection process.

### Data Extraction and Quality Assessment

Two authors extracted data independently and stored them in The Microsoft Excel spreadsheet. Data is then discussed for an agreement. Name of authors, year of publication, country, type of studies, study period, number of patients, comparators, and target conditions was collected. The NOS / Newcastle-Ottawa Scale will be used by authors to assess the quality of the cohort and case-control study, and The Joanna Briggs Institute (JBI) Critical Appraisal Checklist for an analytical cross-sectional study.^11^ Two authors performed the assessment and the results were discussed with the first author.

### Meta-Analysis Outcome

The main outcome of interest was Covid-19 mortality and severity. Covid-19 severity is defined as either ICU admission, ARDS, or need for mechanical ventilation. Covid-19 infection and hospitalization were decided as secondary outcomes.

### Data Analysis & Synthesis

We performed data analysis mainly using Review Manager 5.4.1 (RevMan 5.4.1) by Cochrane collaboration.^12^ Additional synthesis if any were needed then performed with STATA-16. We synthesized the dichotomous outcome from each study with an odds ratio (OR). The random-effects model (DerSimonian and Laird) was used to present pooled OR with 95% CI (Confidence Interval) and the result of overall effect (*p*). We addressed the presence of heterogeneity with *I*^*2*^ as 0% to 40%: might not be important; 30% to 60%: may represent moderate heterogeneity; 50% to 90%: may represent substantial heterogeneity; 75% to 100%: considerable heterogeneity.^13^ We performed subgroup analysis by age, gender, other comorbidities status, cancer type, cancer stage, presence of metastatic disease, and active cancer treatment. Sensitivity analysis would be performed by dividing multi-center/single-center studies and removing/including the latest study period if concerns were raised of patients population duplication thus we could present robust pooled evidence.^14^

## RESULTS

A total of 51 studies involving the Covid-19 positive population, among them are 1991 gynecologic cancer patients, 221465 non-cancer patients, and 28138 other cancer type patients. 3717078 cancer patients were found to be Covid-19 free. Study selection and summary of included studies were presented in **Figure 1** and **Table S1**. The risk of bias in each study was shown in **Fig. S1** and **Fig. S2**.

**Figure 1.**
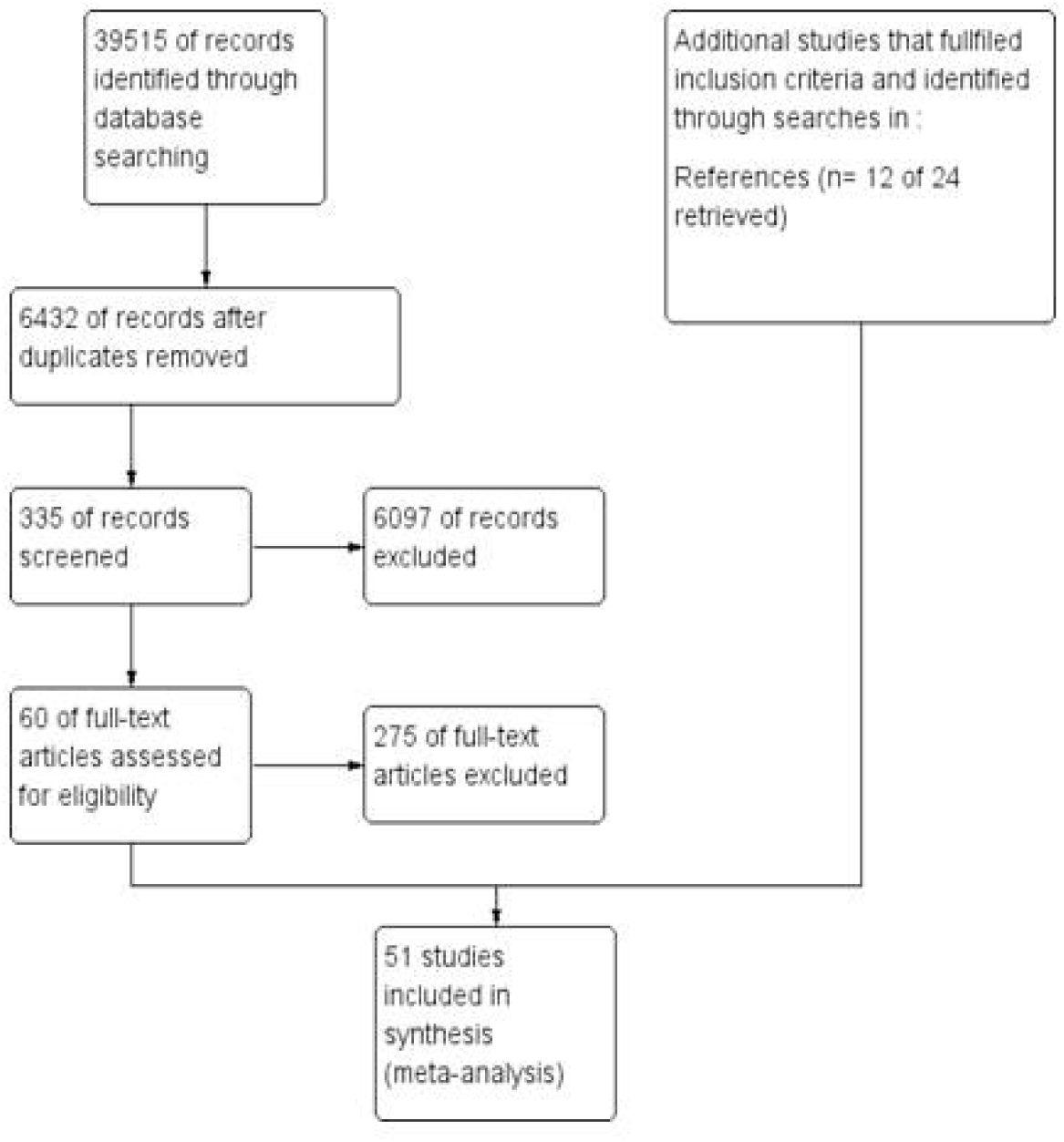
Study Flow Diagram.

### Gynecologic Cancer VS Other Cancer

Covid-19 infection was equivalent between gynecologic cancer and other cancer gathered from 8 studies (OR 1.02, CI 0.84-1.22, *p* 0.87, *I*^*2*^ 57%) **Fig. S3**.^33,39,50,51,55,56^ Gynecologic cancer had fewer Covid-19 associated death compared to other cancer according to 30 studies (OR 0.82, CI 0.71-0.94, *p* 0.006, *I*^*2*^ 0%) **Figure 2**.^18-20,24-28,30,32,37,39-42,45,46,48,50,52-55,57,58,60^ Covid-19 associated severity was not significant from 6 studies between gynecologic cancer and other cancer (OR 0.56, CI 0.30-1.03, *p* 0.06, *I*^*2*^ 0%) **Fig. S4**.^24,25,32,53,54,60^ Data from 2 studies also showed non significance from Covid-19 hospitalization in gynecologic cancer patients than other cancer (OR 0.73, CI 0.50-1.06, *p* 0.10, *I*^*2*^ 82%) **Fig. S5**.^30,50^

**Figure 2.**
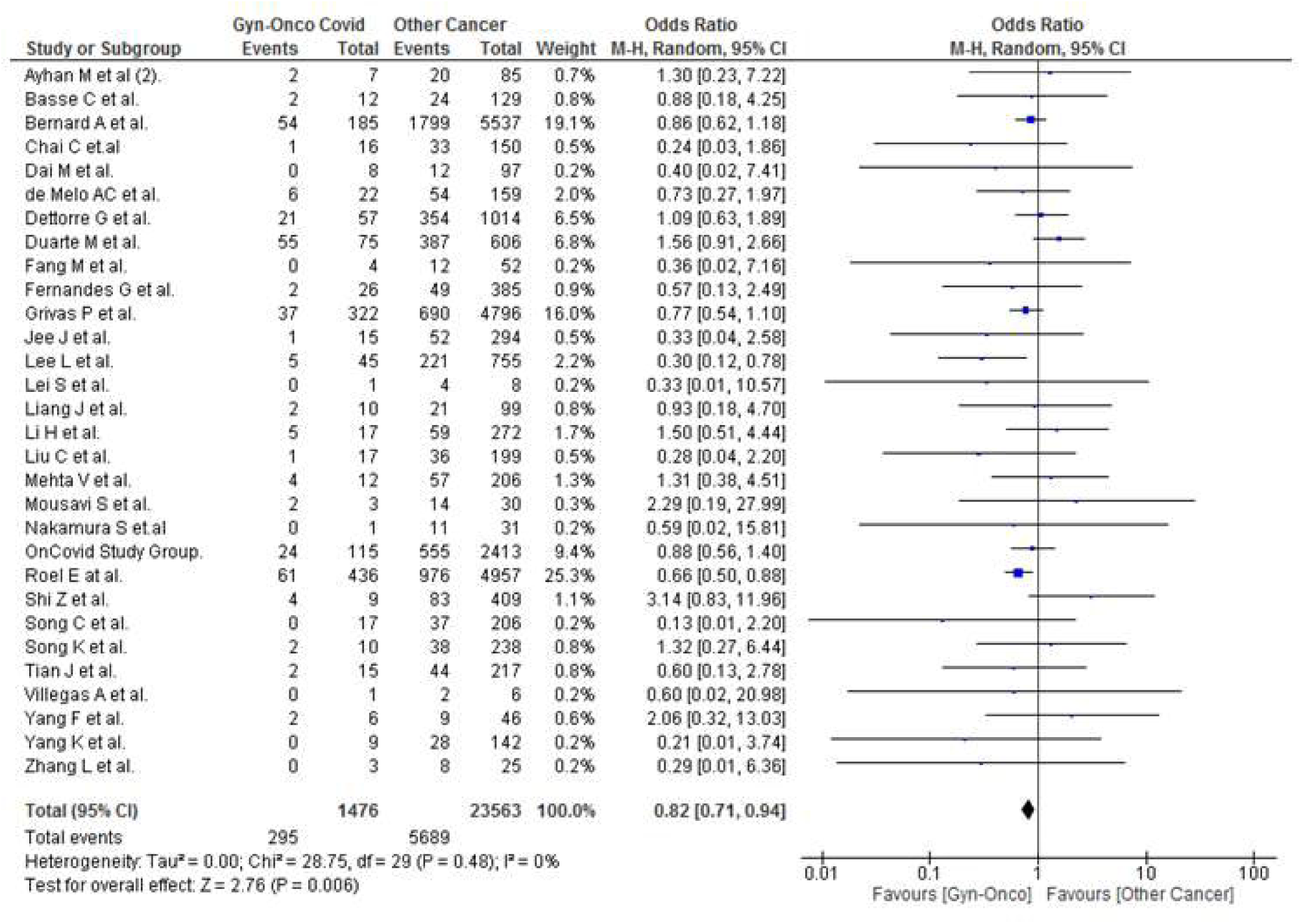
Gynecologic cancer VS Other Cancer, Covid-19 Death.

### Gynecologic Cancer VS Non-Cancer

Covid-19 infection among gynecologic cancer and non cancer population was not significant from 6 studies (OR 1.55, CI 0.81-2.95, *p* 0.18, *I*^*2*^ 90%) **Fig. S6**.^35,39,50,56,59^ Data from 11 studies revealed death from Covid-19 was higher in gynecologic cancer than non cancer patients (OR 2.98, CI 2.23-3.98, *p* <0.0001, *I*^*2*^ 30%) **Figure 3**.^18,20,24,27,38,39,42,50,54^ However, severe Covid-19 was not significant in gynecologic cancer than non cancer patients from 2 studies (OR 1.85, CI 0.77-4.44, *p* 0.17, *I*^*2*^ 0%) **Fig. S7**.^24,54^

**Figure 3.**
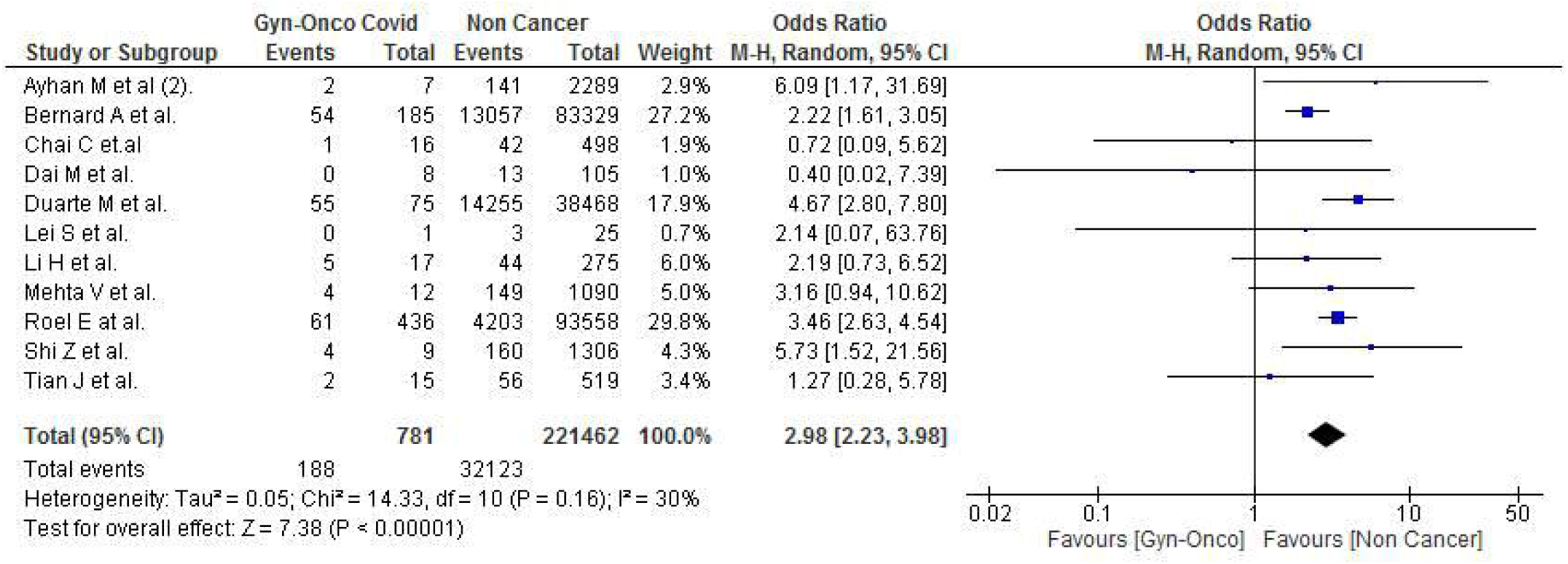
Gynecologic cancer VS Non-Cancer, Covid-19 Death.

### Gynecologic Cancer VS Non-Covid

Data represented from 5 studies revealed that gynecologic cancer patients were experiencing higher Covid-19 associated death in comparison to other cancer patients without Covid-19 infection (OR 11.83, CI 8.20-17.07, *p* <0.0001, *I*^*2*^ 5%) **Figure 4**.^16,39,44,50^

**Figure 4.**
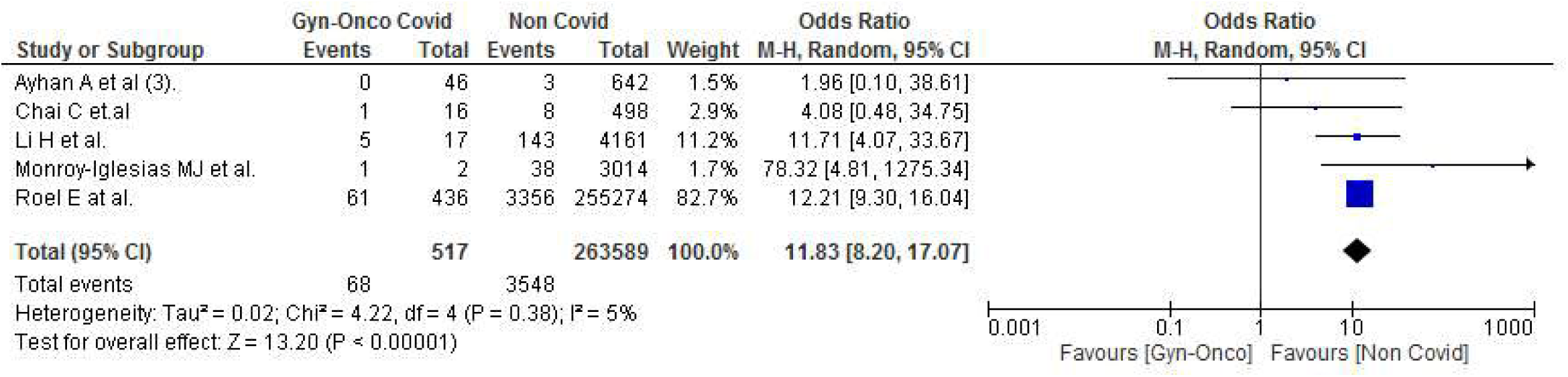
Gynecologic cancer with Covid-19 VS Other cancer non-covid, Covid-19 Death.

### Cancer Treatment Group

We analysed effect of active cancer treatment comprising SACT(Systemic anti cancer therapy), radiotherapy, cancer surgery, and hormonal therapy. Data from 9 studies showed among those who receive active cancer treatment, Covid-19 infection was not significant in gynecologic cancer patients compared to other cancer (OR 0.75, CI 0.55-1.02, *p* 0.07, *I*^*2*^ 0%) **Fig. S8**.^15,17,23,29,31,34,43,47,61^ Covid-19 death was not significant among cancer treatment between gynecologic cancer and other cancer gathered from 9 studies (OR 0.86, CI 0.41-1.78, *p* 0.68, *I*^*2*^ 0%) **Fig. S9**.^15,21,24,25,32,38,44,47,49^ Severe Covid-19 was not significant in gynecologic cancer than other cancer, both who were receiving active cancer treatment according to 6 studies (OR 0.63, CI 0.18-2.25, *p* 0.48, *I*^*2*^ 18%) **Fig. S10**.^24,25,32,44,47,60^ According to 5 studies the Covid-19 death was comparable in gynecologic cancer with active cancer treatment compared to who were not receiving cancer treatment (OR 1.06, CI 0.57-1.98, *p* 0.86, *I*^*2*^ 0%) **Fig. S11**.^22,24,25,32,36^ Lastly, 5 studies showed severity from Covid-19 was not significant in gynecologic cancer who had active cancer treatment than who had none (OR 0.45, CI 0.17-1.20, *p* 0.11, *I*^*2*^ 26%) **Fig. S12**.^24,25,32,36,60^

### Cancer Stage and Metastatic Cancer

2 studies were available for cancer stage analysis.^24,25^ Overall adverse Covid-19 events (infection/hospitalization/severity/death) showed no significance between stage I-II gynecologic cancer against stage III-IV other cancer, stage III-IV gynecologic cancer against stage I-II other cancer, and among all cancer patients who had stage III-IV cancer (OR 0.78, CI 0.04-16.18, *p* 0.88, *I*^*2*^ 67%), (OR 0.48, CI 0.15-1.53, *p* 0.21, *I*^*2*^ 0%), (OR 0.59, CI 0.22-1.58, *p* 0.29, *I*^*2*^ 0%) respectively **Fig. S13-S15**. 3 studies also showed no significance on Covid-19 adverse events between stage III-IV and I-II gynecologic cancer (OR 0.72, CI 0.39-1.33, *p* 0.29, *I*^*2*^ 0%) **Fig. S16**.^24,25,36^

3 studies provided data on metastatic status.^20,25,39^ Gynecologic cancer with metastasis was having increased Covid-19 death than those with localized cancer (OR 1.53, CI 1.06-2.21, *p* 0.02, *I*^*2*^ 0%) **Figure 5**. Contrary, among those who had metastatic diseases, Covid-19 death was not significant between gynecologic cancer compared to other cancer (OR 0.77, CI 0.54-1.11, *p* 0.17, *I*^*2*^ 0%) **Fig. S17**.

**Figure 5.**
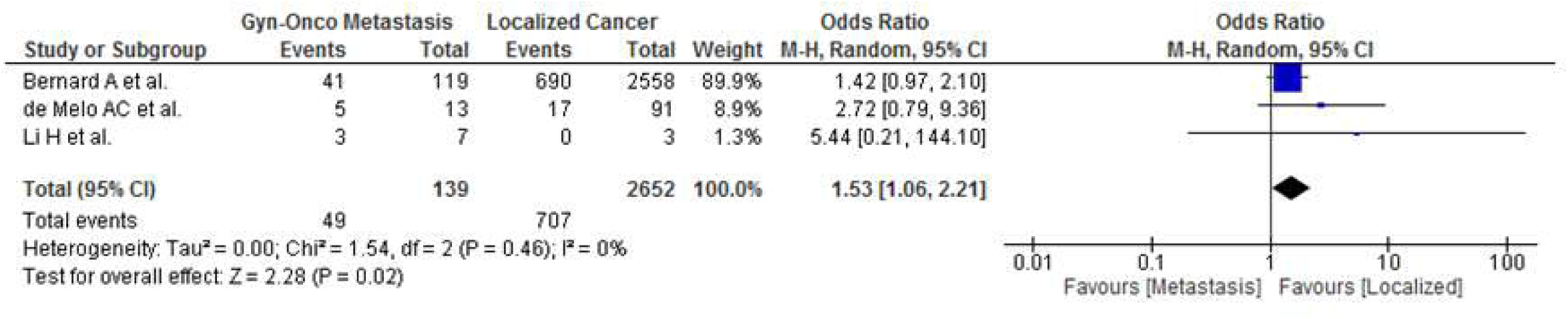
The covid-19 death, Gynecologic cancer with metastasis VS no metastasis.

### Gynecologic cancer VS Lung Cancer

13 studies provided data on Covid-19 infectivity, infection was not significant in gynecologic cancer than lung cancer (OR 0.86, CI 0.61-1.20, *p* 0.37, *I*^*2*^ 73%) **Fig. S18**.^15,17,23,29,33,39,43,50,51,56,61^ Data from 30 studies revealed that gynecologic cancer had fewer Covid-19 death than lung cancer patients (OR 0.52, CI 0.44-.062, *p* <0.0001, *I*^*2*^ 0%) **Figure 6A**.^15,18-21,24-28,30,32,37,39,40-42,45,46,48-50,52-54,57,58^ Data from 6 studies showed that gynecologic cancer was having less severity from Covid-19 than lung cancer (OR 0.36, CI 0.16-0.80, *p* 0.01, *I*^*2*^ 0%) **Figure 6B**.^24,25,32,53,54,60^ Lastly, 2 studies reported fewer hospitalization associated with Covid-19 in gynecologic cancer than lung cancer (OR 0.54, CI 0.40-0.73, *p* <0.0001, *I*^*2*^ 0%) **Figure 6C**.^17,30^

**Figure 6A.**
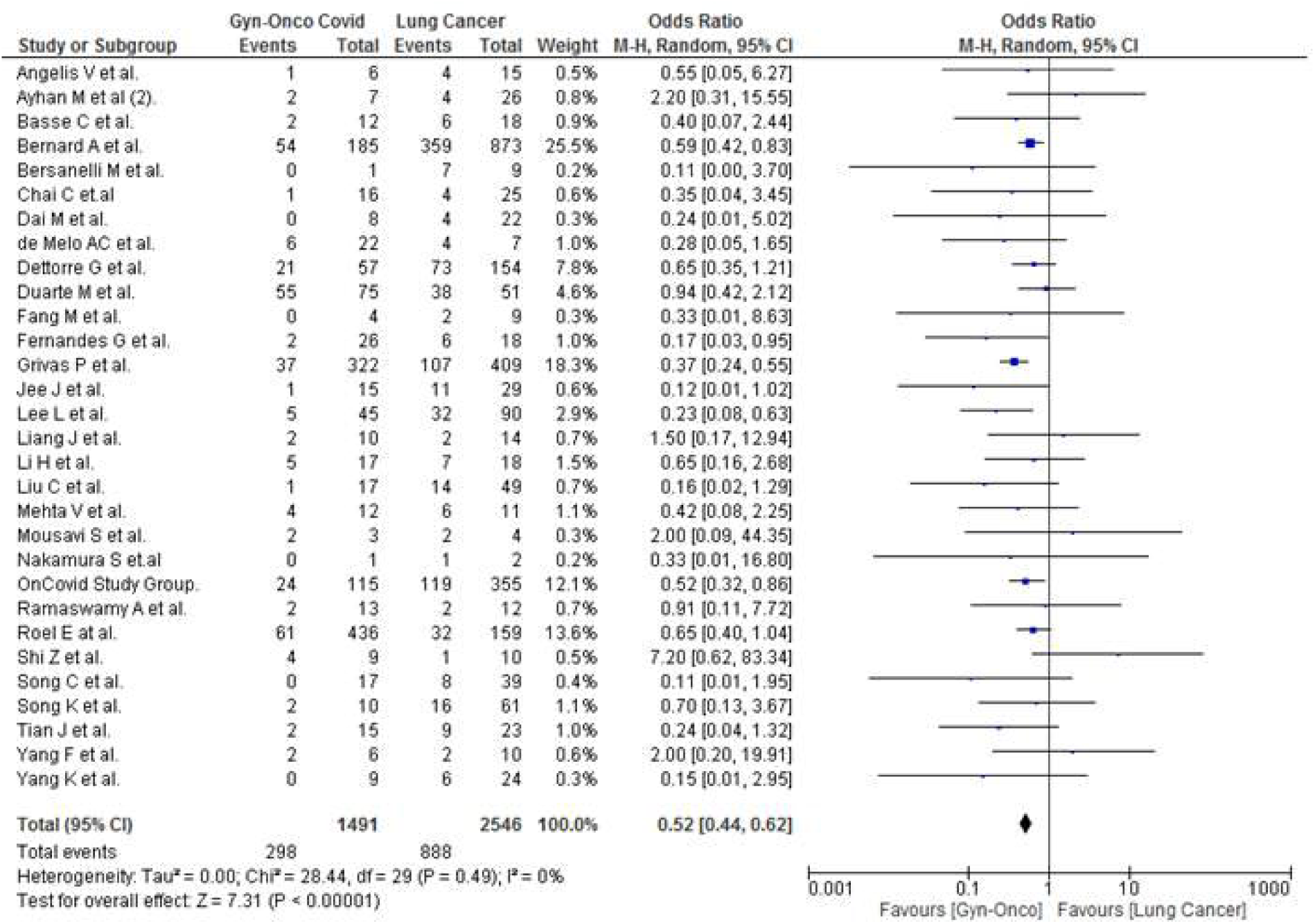
Gynecologic cancer VS Lung Cancer, covid-19 death.

**Figure 6B.**
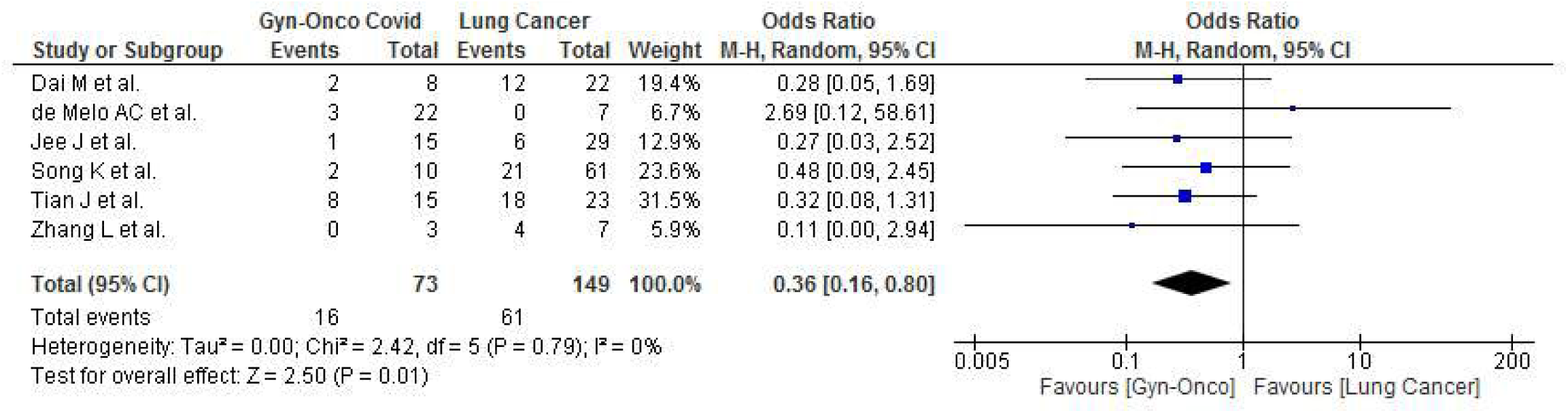
Gynecologic cancer VS Lung Cancer, severe Covid-19.

**Figure 6C.**
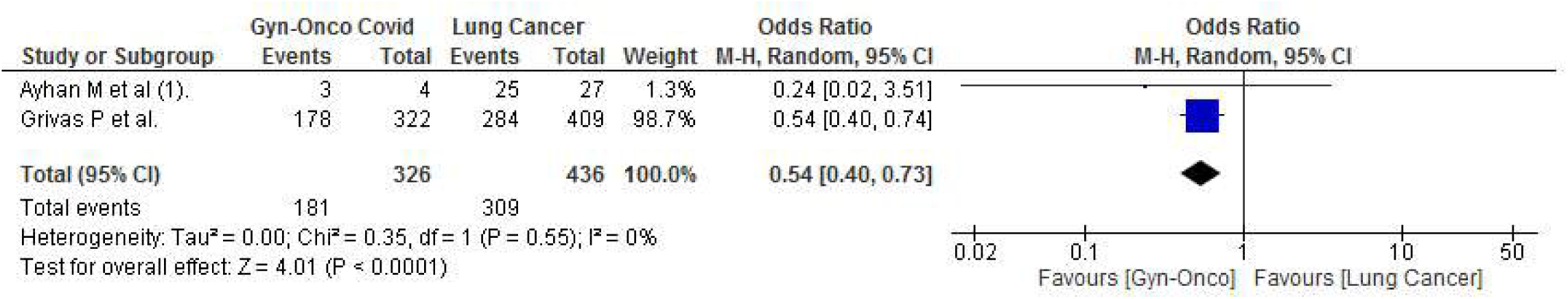
Gynecologic cancer VS Lung Cancer, covid-19 hospitalization.

### Gynecologic cancer VS Breast Cancer

Data from 13 studies showed gynecologic cancer and breast cancer were equivalent on having Covid-19 infection (OR 1.05, CI 0.94-1.17, *p* 0.37, *I*^*2*^ 7%) **Fig. S19**.^15,17,29,33,39,43,47,50,51,56,61^ Interestingly from 25 studies, gynecologic cancer was experiencing higher Covid-19 death compared to breast cancer patients (OR 1.50, CI 1.20-1.88, *p* 0.0004, *I*^*2*^ 19%) **Figure 7A**.^15,18-20,25-28,30,32,37,39-42,45,48-50,53,54,57,58^ Covid-19 severity was not significant from 7 studies between gynecologic cancer and breast cancer (OR 0.83, CI 0.40-1.72, *p* 0.62, *I*^*2*^ 0%) **Fig. S20**.^24,25,32,47,53,54,60^ Lastly, data from 2 studies showed gynecologic cancer was experiencing higher hospitalization from Covid-19 compared to breast cancer (OR 1.52, CI 1.18-1.96, *p* 0.001, *I*^*2*^ 0%) **Figure 7B**.^17,30^

**Figure 7A.**
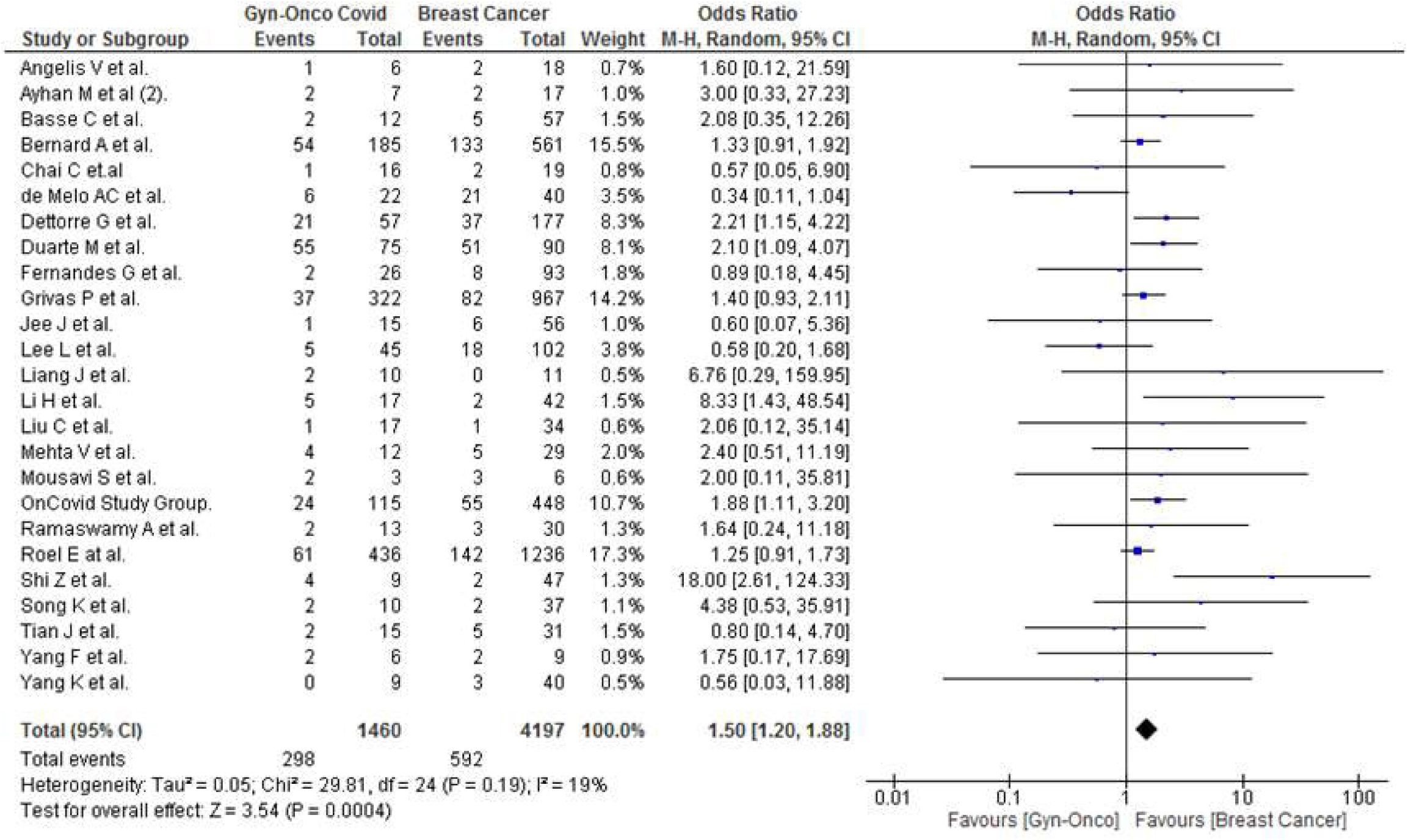
Gynecologic cancer VS Breast Cancer, covid-19 death.

**Figure 7B.**
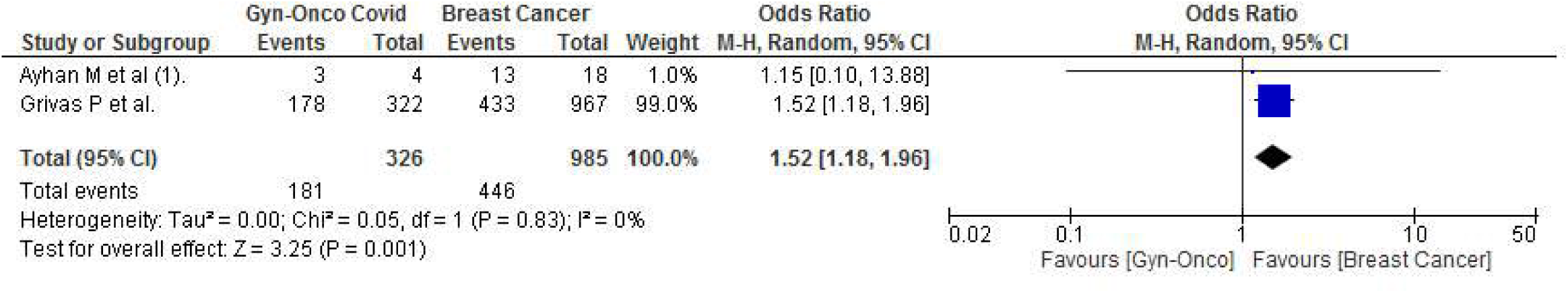
Gynecologic cancer VS Breast Cancer, covid-19 hospitalization.

### Gynecologic cancer VS Hematologic Cancer

Data available from 8 studies revealed gynecologic cancer was having less infection of Covid-19 compared to hematologic cancer patients (OR 0.71, CI 0.56-0.90, *p* 0.005, *I*^*2*^ 68%) **Figure 8A**.^15,33,39,50,51,56^ Data also showed that gynecologic cancer was experiencing fewer Covid-19 death compared to hematologic cancer from 24 studies (OR 0.63, CI 0.47-0.83, *p* 0.001, *I*^*2*^ 46%) **Figure 8B**.^15,19,20,24-28,30,32,37,39,40,42,46,48-50,52,54,58^ Lastly, 4 studies also showed that gynecologic cancer was having less severity from Covid-19 compared to hematologic cancer (OR 0.26, CI 0.10-0.67, *p* 0.005, *I*^*2*^ 0%) **Figure 8C**.^24,25,32,54^

**Figure 8A.**
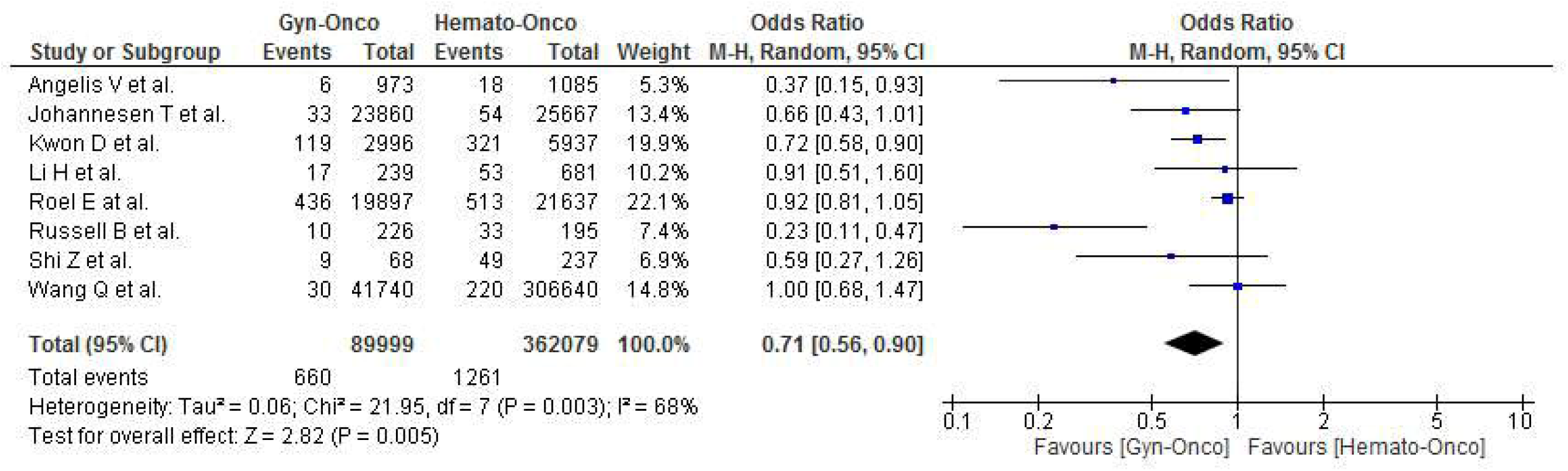
Gynecologic cancer VS Hematologic Cancer, covid-19 infection.

**Figure 8B.**
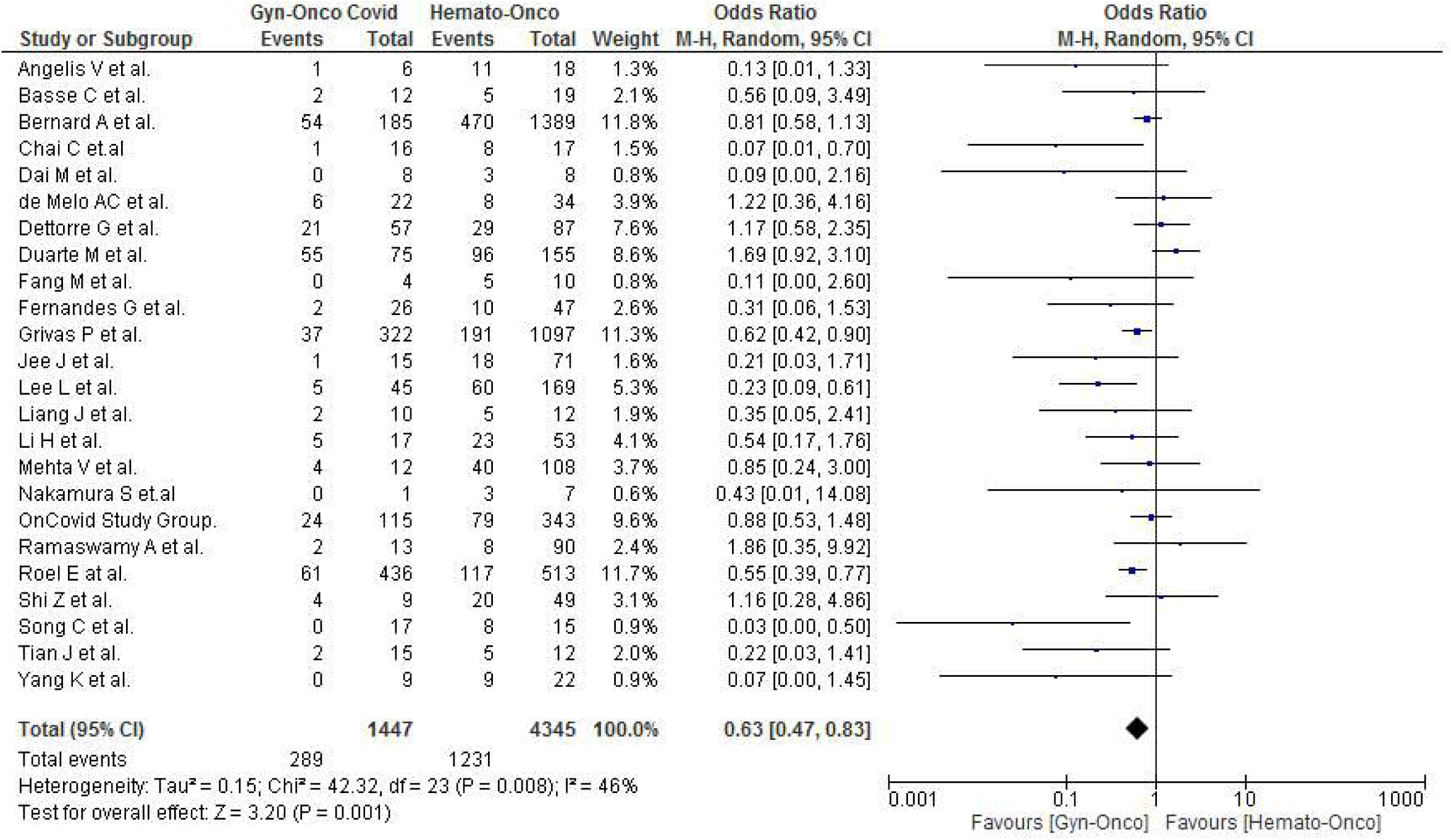
Gynecologic cancer VS Hematologic Cancer, covid-19 death.

**Figure 8C.**
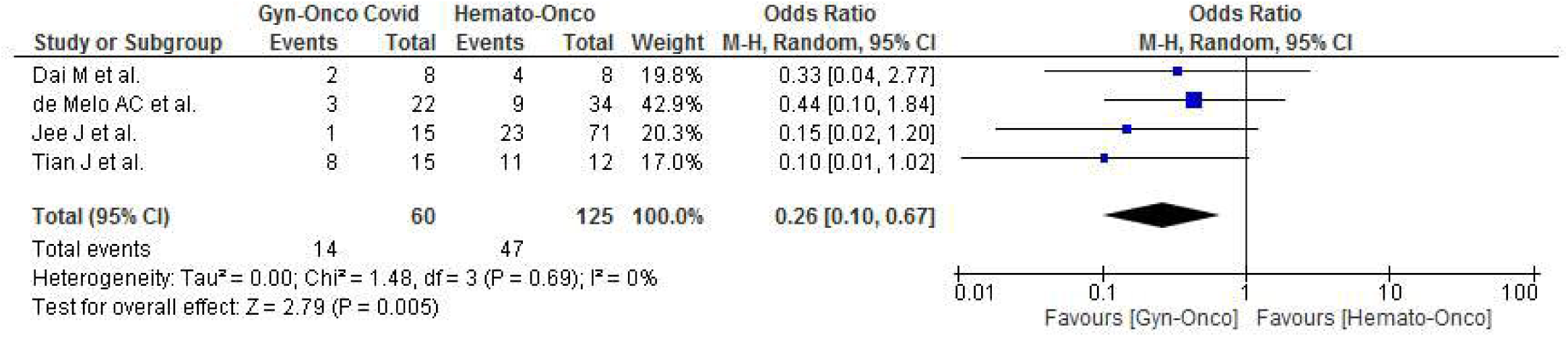
Gynecologic cancer VS Hematologic Cancer, severe Covid-19.

### Gynecologic Cancer VS Men

Based on 10 studies available for synthesis, there was no significance on Covid-19 infection between gynecologic cancer population and men with cancer (OR 0.58, CI 0.27-1.22, *p* 0.15, *I*^*2*^ 94%) **Fig. S21**.^17,23,29,39,43,51,56,61^ Compared to men with cancer and gynecologic cancer patients, the Covid-19 associated death retrieved from 23 studies showed no significance (OR 0.75, CI 0.54-1.05, *p* 0.09, *I*^*2*^ 23%) **Fig. S22**.^15,18,21,24,25,27,28,30,32,37,39-42,46,49,52,53,57,58^

According to 6 studies, severe Covid-19 was higher in men with cancer compared to gynecologic cancer patients (OR 0.47, CI 0.25-0.88, *p* 0.02, *I*^*2*^ 0%) **Figure 9A**.^24,25,32,53,54,60^ Hospitalization from Covid-19 was also higher in men with cancer compared to gynecologic cancer patients synthesized from 2 studies (OR 0.71, CI 0.56-0.89, *p* 0.004, *I*^*2*^ 0%) **Figure 9B**.^17,30^

**Figure 9A.**
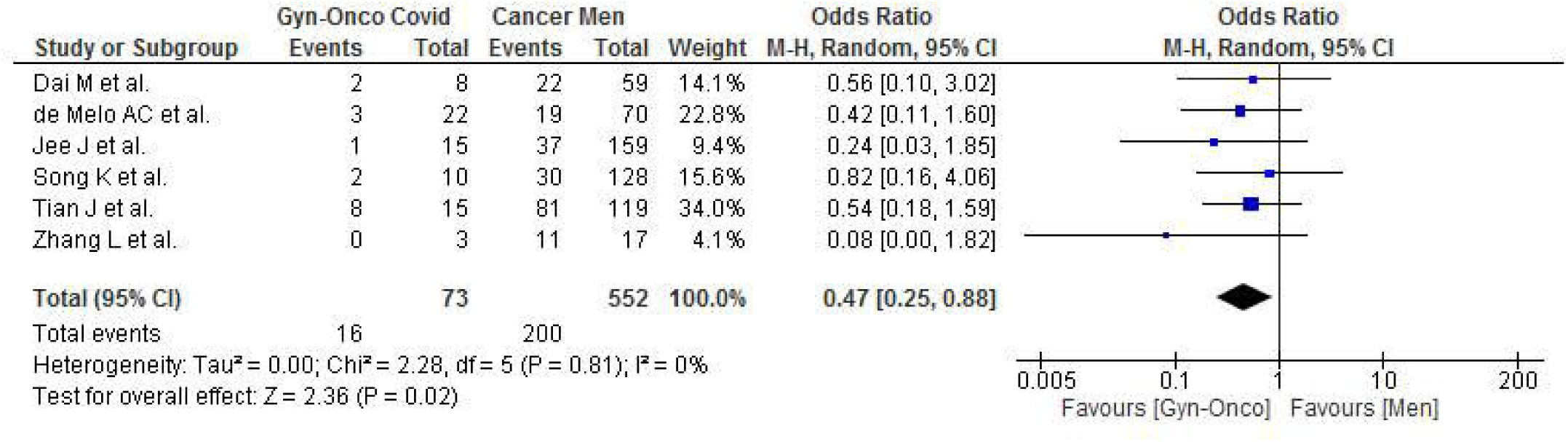
Gynecologic cancer patients VS Men with cancer, severe Covid-19.

**Figure 9B.**
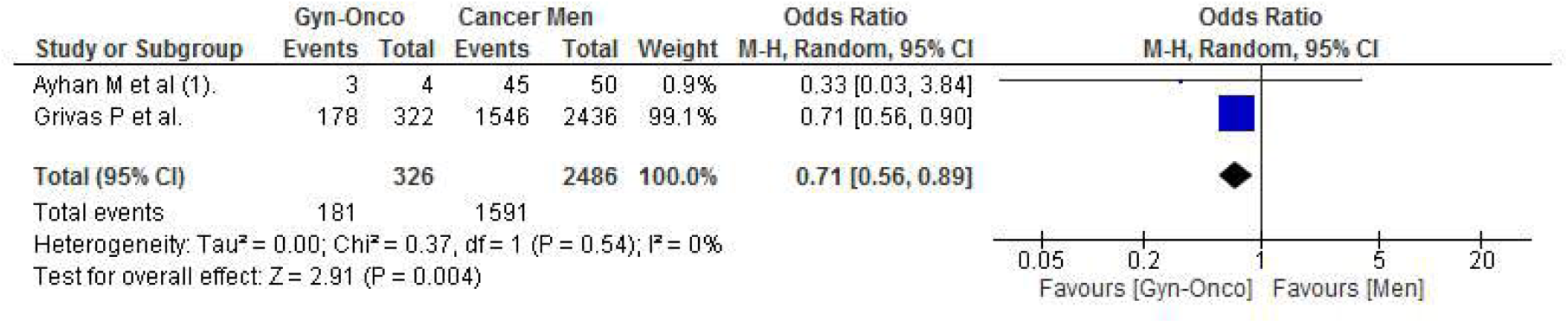
Gynecologic cancer patients VS Men with cancer, covid-19 hospitalization.

### Age Stratification

Data from 4 studies showed that among the gynecologic cancer population, those who were >65 compared to <65 years of age had comparable overall adverse Covid-19 outcomes (infection/hospitalization/severity/death), (OR 1.13, CI 0.48-2.62, *p* 0.78, *I*^*2*^ 14%) **Fig. S23**.^16,22,24,25^ We performed a pairwise comparison of gynecologic cancer with <65 years old against other cancer with >65 years old, and gynecologic cancer with >65 years old against other cancer with <65 years old.^24,25,60^ Covid-19 adverse outcome was seen lower in <65 years old gynecologic cancer than >65 years old other cancer population (OR 0.16, CI 0.06-0.47, *p* 0.0007, *I*^*2*^ 0%) **Figure 10**. Contrary, there was an equivalent Covid-19 adverse outcome between gynecologic cancer with >65 years old and other cancer with <65 years old (OR 1.08, CI 0.36-3.26, *p* 0.89, *I*^*2*^ 0%) **Fig. S24**.

**Figure 10.**
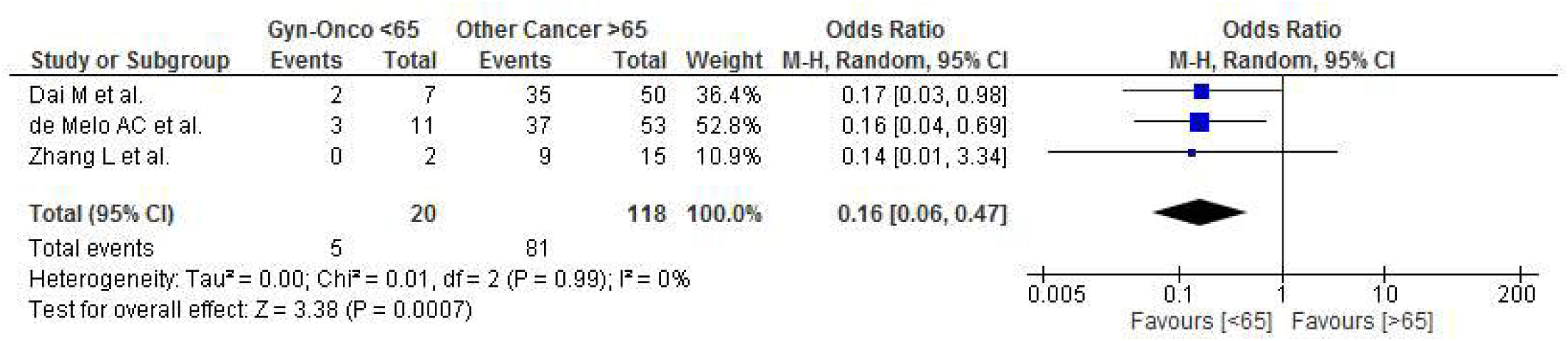
The covid-19 adverse outcome, <65 years old gynecologic cancer VS >65 years old other cancer population.

### Comorbidities

Cancer is a comorbidity, aside from that we tried to analyze other comorbidities (Hypertension, diabetes, cardiovascular disease, pulmonary disease, renal disease, liver disease, immune disease, metabolic-endocrine disease) present within the cancer population. Among those with comorbidities, gynecologic cancer had few adverse Covid-19 outcomes than other cancer populations according to 4 studies (OR 0.31, CI 0.12-082, *p* 0.02, *I*^*2*^ 0%) **Figure 11**.^21,24,25,60^ 5 studies showed there was no significant adverse Covid-19 outcome between gynecologic cancer patients with comorbidities against no comorbidities (OR 2.34, CI 0.59-9.79, *p* 0.24, *I*^*2*^ 79%) **Fig. S25**.^16,22,24,25,36^ Gynecologic cancer without comorbidities against other cancer with comorbidities had no significant adverse Covid-19 outcome, according to 3 studies (OR 0.29, CI 0.04-2.22, *p* 0.23, *I*^*2*^ 56%) **Fig. S26**. ^24,25,60^ Gynecologic cancer with comorbidities against other cancer without comorbidities also showed no significant adverse Covid-19 outcome, according to 4 studies (OR 0.61, CI 0.22-1.72, *p* 0.35, *I*^*2*^ 0%) **Fig. S27**.^21,24,25,60^

**Figure 11.**
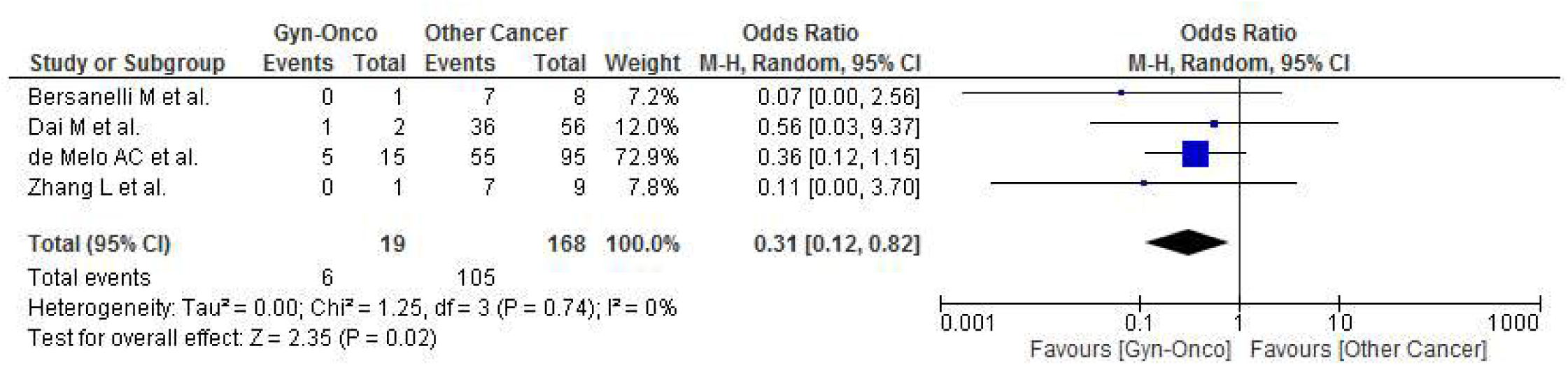
The covid-19 adverse outcome with comorbidities, Gynecologic cancer VS Other cancer.

### Sensitivity Analysis

We performed sensitivity analysis by reproducing each outcome synthesis to pre-specified single center to multi-center studies, furthermore excluding overlapped study periods associated with its study centers thus only one center with the most recent study period was included in **Table S2**. After exclusion of 3 studies, a difference of significance was found in severe Coivd-19 between gynecologic cancer and cancer men population (OR 0.47, CI 0.19-1.17, *p* 0.10, *I*^*2*^ 0%)^25,32,53^ Aside from that, the remainder of calculated OR from reproducing each outcome synthesis by exclusion were within good accordance.

### Publication Bias

We found no publication bias within our included studies though at first, we identified an asymmetrical funnel plot; it was caused solely by heterogeneity nonetheless (**Fig. S28-31)**. After subgroup identification, the funnel plow was corrected and the calculated Egger & Begg’s test for overall Covid death, severity, and hospitalization were *p* 0.15 and *p* 1.6. For data associated with Covid-19 infection, the values were *p* 0.17 and *p* 1.87.

## DISCUSSION

We believe this is the first comprehensive meta-analysis with a large population regarding the outcome of Covid-19 on the gynecologic cancer population. With the 1991 Covid-19 positive gynecologic cancer, we hope we provide new insight into how the global pandemic is affecting practice and services affecting gynecologic cancer. Several meta-analyses showed the prevalence of cancer with Covid-19 infection was 2-4%, Covid-19 mortality also higher in the cancer patients cohort.^5-7,62-66^ In this meta-analysis gynecologic cancer patients are at increased risk of Covid-19 death compared to the non-cancer population (OR 2.98, CI 2.23-3.98, *p* <0.0001, *I*^*2*^ 30%), most studies also support this finding by providing evidence of greater Covid-19 adverse outcome in cancer patients.^5-7,62-66^ Contrary to the “*N3C*” multicenter study from the United States, our result present a significant increased of death in gynecologic cancer with Covid-19 than other cancer types without Covid-19 (OR 11.83, CI 8.20-17.07, *p* <0.0001, *I*^*2*^ 5%).^67^ Our finding shows gynecologic cancer with metastatic disease has an increased Covid-19 death compared to those whose cancer is localized (OR 1.53, CI 1.06-2.21, *p* 0.02, *I*^*2*^ 0%), most studies also report identical outcomes to ours.^66,68,69^ Our analysis also shows gynecologic cancer is associated with higher Covid-19 death and hospitalization compared to breast cancer patients (OR 1.50, CI 1.20-1.88, *p* 0.0004, *I*^*2*^ 19%), (OR 1.52, CI 1.18-1.96, *p* 0.001, *I*^*2*^ 0%) respectively. Other meta-analyses, as well as studies done by “*CCC19*” and the “*N3C*” also supported this finding.^63,67,68^ Our analysis present that gynecologic cancer patients have lower Covid-19 death compared to overall other cancer types (OR 0.82, CI 0.71-0.94, *p* 0.006, *I*^*2*^ 0%), further analysis shows that gynecologic cancer patients with Covid-19 have fewer adverse outcome compared to Covid-19 lung and hematologic cancer. Our findings are (OR 0.52, CI 0.44-.062, *p* <0.0001, *I*^*2*^ 0%), (OR 0.36, CI 0.16-0.80, *p* 0.01, *I*^*2*^ 0%), (OR 0.54, CI 0.40-0.73, *p* <0.0001, *I*^*2*^ 0%) for Covid-19 associated death, severity, and hospitalization versus lung cancer respectively. Hematologic cancer (OR 0.71, CI 0.56-0.90, *p* 0.005, *I*^*2*^ 68%), (OR 0.63, CI 0.47-0.83, *p* 0.001, *I*^*2*^ 46%), (OR 0.26, CI 0.10-0.67, *p* 0.005, *I*^*2*^ 0%) for Covid-19 infectivity, death, and severity respectively. The “*TERAVOLT*” study and the one conducted by *Luo et al*. also support our finding of a high Covid-19 associated adverse outcomes among lung cancer patients.^70,71^ Other meta-analyses show lung cancer with Covid-19 has a 32.9% case fatality rate (378 lung cancer), compared to the non-lung cancer population the Covid-19 death among lung cancer is also higher (92 lung cancer, 554 control, OR 1.83, *p* 0.05), (78 lung cancer, 482 control, RR 1.46, *p* 0.7).^5,63,64^ Lastly, most studies also support our findings on the increased Covid-19 adverse outcome in the hematologic cancer population, as their results are 34.2% case fatality rate (480 hematologic cancer), (120 hematologic cancer, 758 control, OR 2.39, *p* 0.02).^63,64,66-69.^ We believe our meta-analysis results correspond to several studies that present the safety of continuing gynecologic cancer care and service during the global pandemic. Safety protocols have been published for gynecologic cancer patients who are seeking treatment and some even recommend the implementation of ERAS (Enhanced Recovery After Surgery).^2,72,73^ Data from the French Society for Pelvic and Gynecological Surgery (SCGP) and the French (FRANCOGYN) Group reveal there are changes in cancer management strategy during the pandemic time and from 181 gynecologic cancer patients, 8 tested positive of Covid-19.^74^ Multicenter study from three New York City hospitals also show a similar result, among 302 gynecologic cancer patients, 117 experienced a COVID-19-related treatment modification, 19 have positive Covid-19 result among them 3 are asymptomatic, 11 are having mild symptoms, 3 are hospitalized, and 2 died.^75^ Lastly, data from the United Kingdom, Turkey, and Italy show that while maintaining gynecologic cancer treatment during the pandemic time the Covid-19 infection rate is found at a low level, 1/289 is Covid-19 positive and 1 post-operative death suspected of Covid-19 (UK), 2/200 is suspected with Covid-19 but neither was positive for COVID-19 on polymerase chain reaction testing (Turkey), and 1/930 is Covid-19 positive (Italy).^76-78^ Meta-analysis shows Covid-19 infection with existing comorbidities such as hypertension (OR 1.95, *p* <0.0001), diabetes (OR 1.97, *p* <0.0001), respiratory disease (OR 2.74, *p* <0.0001), cardiovascular disease (OR 3.05, *p* <0.0001), cerebrovascular disease (OR 4.78, *p* <0.0001), kidney disease (OR 4.90, *p* <0.0001), and cancer (OR 1.89, *p* <0.0001) increase the risk of mortality.^79^ Our analyzed population comprises cancer as the main comorbidity, however with comorbidities other than cancer, our study shows that the gynecologic cancer population with additional comorbidities has fewer adverse events than other cancer with comorbidities (OR 0.31, CI 0.12-082, *p* 0.02, *I*^*2*^ 0%). Meta-analyses prove that men have increased Covid-19 severity and mortality.^79,80^ Our findings correspond by showing that severity and hospitalization from Covid-19 were higher in men with cancer compared to gynecologic cancer patients (OR 0.47, CI 0.25-0.88, *p* 0.02, *I*^*2*^ 0%), (OR 0.71, CI 0.56-0.89, *p* 0.004, *I*^*2*^ 0%) respectively. Age thresholds above 50 and 60 years old have an effect on Covid-19 mortality.^79,81^ In our study Covid-19 adverse outcome was lower in <65 years old gynecologic cancer than <65 years old other cancer patients (OR 0.16, CI 0.06-0.47, *p* 0.0007, *I*^*2*^ 0%). Meta-analysis on Covid-19 with active cancer treatment shows that cancer surgery (OR 1.14, *p* <0.01), chemotherapy (OR 1.60, <0.01), and overall cancer treatment type (OR 1.16, *p* <0.01) have a higher risk of death.^82^ However in our study Covid-19 death is equivalent in gynecologic cancer with active cancer treatment compared to those who are not receiving cancer treatment (OR 1.06, CI 0.57-1.98, *p* 0.86, *I*^*2*^ 0%).

We hope these findings will be useful among gynecologist-oncologists in cancer centers or tertiary cancer referral centers who provide care to gynecologic cancer patients during the ongoing Covid-19 pandemic.

### Limitations

In several data syntheses with the statistically nonsignificant value, we analyze few data regarding severity, hospitalization, age, cancer stage/metastatic status, other comorbidities aside from cancer, and cancer treatment type due to limited data, however those aforementioned are well represented and distributed through other synthesis based on the patient’s characteristics available in **Table 1**.

## Supporting information

Reporting guideline

## Data Availability

The report of the following are publicly available and they can be found in the supplementary material.

## Acknowledgments

We thank the staff of Gynecology Oncology (Sanglah Hospital, Faculty of Medicine, Udayana University, Denpasar, Bali, Indonesia), staff of Reproductive Endocrinology and Infertility (Morula IVF), (School of Medicine and Health Sciences, Atmajaya Catholic University of Indonesia, Jakarta, Indonesia), and staff of Department of Obstetrics and Gynecology (UKI Hospital, Faculty of Medicine, Christian University of Indonesia, Jakarta, Indonesia) to make this research collaboration possible.

## Conflict of Interest

None declared.

## Availability of data, code, and other materials

**Figure 1**. Study Flow Diagram.

**Figure 2**. Gynecologic cancer VS Other Cancer, Covid-19 Death.

**Figure 3**. Gynecologic cancer VS Non-Cancer, Covid-19 Death.

**Figure 4**. Gynecologic cancer with Covid-19 VS Other cancer non-covid, Covid-19 Death.

**Figure 5**. The covid-19 death, Gynecologic cancer with metastasis VS no metastasis.

**Figure 6**. Gynecologic cancer VS Lung Cancer, **(A)** Covid-19 death. **(B)** Severe Covid-19. **(C)** Covid-19 hospitalization.

**Figure 7**. Gynecologic cancer VS Breast Cancer, **(A)** Covid-19 death, **(B)** Covid-19 hospitalization.

**Figure 8**. Gynecologic cancer VS Hematologic Cancer, **(A)** Covid-19 infection. **(B)** Covid-19 death. **(C)** Severe Covid-19.

**Figure 9**. Gynecologic cancer patients VS Men with cancer. **(A)** Severe Covid-19, **(B)** Covid-19 hospitalization.

**Figure 10**. The covid-19 adverse outcome, <65 years old gynecologic cancer VS >65 years old other cancer population.

**Figure 11**. The covid-19 adverse outcome with comorbidities, Gynecologic cancer VS Other cancer.

## Notes

### Competing Interest Statement

The authors have declared no competing interest.

### Clinical Protocols

https://www.crd.york.ac.uk/prospero/display_record.php?RecordID=256557

### Funding Statement

This study did not receive any funding

### Summary of Updates

Additional figures and data synthesis.

