## Supplemental Table 1. for "The Outcome of Gynecologic Cancer Patients With Covid-19 Infection: A Systematic Review And Meta-Analysis"

**Table S1**. Characteristics of included studies.

| Author | Location | Type of Study | Time of Study | Publication Year | Non Cancer Covid Patients | Gynecology Oncology Covid Patients | Other Oncology Covid Patients | Cancer Non Covid Patients | Gender* | Cancer Stage* | Comorbidities* | Cancer Treatment* | Age* | Outcome |
| --- | --- | --- | --- | --- | --- | --- | --- | --- | --- | --- | --- | --- | --- | --- |
| Angelis V et al.^15^ | United Kingdom | Multi center,prospective cohort | March-April 2020 | 2020 | NA | 6 | 107 (Lung 15,Breast 18,Hematological 18) | 13376 (Gynecological 967) | Male 63,Female 50 | NA | Hypertension 39,Diabetes 18,Ischemic heart disease 13,COPD 6 | SACT 85,Radiotherapy 11 | Median 66,IQR: 54-69,range 21-91 | Covid Infection & Covid Death |
| Ayhan A et al (3).^16^ | Turkey | Multi center,retrospective cohort | March-April 2020 | 2020 | NA | 46 | NA | 642 (Gynecological) | Female 688 | NA | Hypertension 29,Diabetes 16,Chronic pulmonary disease 11,Coronary heart disease 6,CKD 1. | Major / Complex Cancer Surgery 688 | <65: 34, >65: 12 | Covid Death |
| Ayhan M et al (1).^17^ | Turkey | Single center, retrospective cohort | March-June 2020 | 2021 | NA | 4 (Ovarian 1,Endometrium 3) | 80 (Lung 27,Breast 18) | 1065 (Ovarian 59, Endometrium 21) | Female 33,Male 51 | I: 2,II: 7,III: 18,IV:57,Metastasis 57,Non-meta 27 | Hypertension 12,Diabetes 16, Coronary artery disease 3,COPD 3,CKD 1 | SACT 84 | Median 61,IQR: 21-84 | Covid Infection & Covid Hospitalization |
| Ayhan M et al (2).^18^ | Turkey | Single center,retrospective cohort | March - May 2020 | 2021 | 2289 | 7 (Cervix 3, Endometrial 2, Ovarian 2) | 85 (Lung 26,Breast 17) | NA | Female 41,Male 51 | Metastasis 53,Non-meta 39 | Hypertension 31,Dibetes 16,COPD 14,Coronary artery disease 13,CKD 4,Chronic liver disease 2,Cerebrovascular disease 2 | SACT 62 | <67: 45, >67: 47 | Covid Death |
| Basse C et al.^19^ | France | Single center,prospective cohort | March 2020 | 2020 | NA | 12 | 129 (Lung 18,Breast 57,Hematological 19) | NA | Female 102,Male 39 | Localized 38,Metastasis 84 | Chronic lung disease 7,Diabetes 24,Hypertension 48,Other heart disease 21,Systemic disease 6 | Surgery 11,Radiotheraphy 13,SACT 120,None 17 | >70: 141 | Covid Death |
| Bernard A et al.^20^ | France | Multi center,retrospective cohort | March-April 2020 | 2021 | 83329 | 185 | 5537 (Lung 873,Breast 561,Hematological 1389 ) | NA | Female 39919, Male 45079 | Metastasis 1775,Non-meta 2558 | Hypertension 28163,Heart failure 6641,Chronic respiratory disease 1334,CKD 6948,Diabetes 16216,COPD 4516,Obesity 8289,Chirosis 673 | NA | With cancer: mean 72,Without cancer: mean 65 | Covid Death |
| Bersanelli M et al.^21^ | Italy | Multi center,prospective cohort | January-April 2020 | 2020 | NA | 1 (Endometrial) | 13 (Lung 9,Breast 1) | 52 | Female 3,Male 10 | IV: 9 | Splenectomy 1,Hypertension 8,HIV 1,Diabetes 1 | ICI 13, ICI+Chemotherapy 1 | <65: 5,>65: 9 | Covid Death |
| Bogani G et al.^22^ | Italy | Single center,retrospective cohort | February-March 2020 | 2020 | NA | 19 (Ovarian 14, Endometrial 3, Cervical 1, Ovarian+Endometrial 1) | NA | 336 (Gynecological) | Female 19 | NA | Cardiovascular disease 5,CKD 1,Hypothyroidism 2,Plummer disease 1 | Surgery 5,SACT 8,Planned treatment 6 | <65: 9,>65: 10 | Covid Death |
| Cavanna L et al.^23^ | Italy | Single center,retrospective cohort | April-June 2020 | 2021 | NA | 0 | 10 (Lung 2) | 250 (Gynecologic cancer 29) | Female 2,Male 8 | NA | NA | SACT 7,Hormonal 1 | Mean 69.2,Range 54-80 | Covid Infection |
| Chai C et al. | China | Multi center,prospective cohort | January-March 2020 | Pre-prints | 498 | 16 (Cervical 9,Ovarian 4,Endometrial 3) | 150 (Lung 25,Breast 19,Hematological 17) | 498 | Female 336,Male 328 | NA | Hypertension 226,Diabetes 128,Hyperlipidemia 109,Heart disease 78,Cerebrovascular disease 22,COPD 36,CKD 14,Chronic liver disease 12 | NA | Median 65,IQR 59-70 |  |
| Dai M et al.^24^ | China | Multi center,prospective cohort | January-February 2020 | 2020 | 105 | 8 (Cervical 6, Ovarian 1, Endometrial 1) | 97 (Lung 22,Breast 11,Hematological 8) | NA | Female 46,Male 59 | I/II: 42,III/IV: 37,Metastasis 17 | Hypertension 160,Cardiovascular disease 51,Diabetes 36,Cerebrovascular disease 26,CKD 28,Chronic liver disease 42 | Surgery 8,SACT 27,Radiotherapy 13 | <65: 54,>65: 51 | Covid Death and Severe Covid |
| de Melo AC et al.^25^ | Brazil | Single center,retrospective cohort | April-May 2020 | 2020 | NA | 22 (Cervical 12, Ovarian 3, Endometrial 5, Vulvar 2) | 159 (Lung 7,Breast 40,Hematological 34) | NA | Female 110, Male 71 | I/II: 27,III/IV: 124,Metastasis 87 | Hypertension 77,diabetes 31,CKD 10,COPD/Asthma 7 | Surgery 12,Radiotheraphy 10,SACT 88,Palliative 32,Hormonal 20 | <60: 89,60-74: 67,>75: 25 | Covid Death and Severe Covid |
| Dettore G et al.^26^ | OnCovid-Europe | Multi center,prospective cohort | February-June 2020 | 2021 | NA | 57 | 1014 (Lung 154,Breast 177,Hematological 87) | NA | Female 231,Male 296 | Localized 173,Metastatic 223 | Hypertension 251,Diabetes 115,Cardiovascular disease 128,Chronic pulmonary disease 80,CKD 62,Cerebrovascular disease 37,Liver impairment 11,Immunosuppression 16 | Ongoing treatment at diagnosis 516,Surgery 510,SACT 319,Radiotherapy 319,Palliative 277 | Mean: 67.9 | Covid Death |
| Duarte M et al.^27^ | Brazil | Multi center,retrospective cohort | January-September 2020 | 2020 | 38468 | 75 (Cervix 47,Uterine 6,Ovaries 22) | 606 (Lung 51,Breast 90,Hematological 155 ) | NA | Female 374,Male 307 | I/II: 106,III/IV: 444 | Heart disease 143,Diabetes 104,Neurologic disease 13,Chronic lung disease 29,Nephropathy 39 | SACT 431 | <65: 441,>65: 240 | Covid Death |
| Fang M et al. | China | Single center,retrospective cohort | February-April 2020 | pre-prints | NA | 4 | 52 (Lung 9,Breast 4,Hematological 10) | NA | Female 24,Male 32 | NA | Hypertension 23,Diabetes 7,Cardiovascular disease 5,Chronic pulmonary disease 1,Chirrosis 2,CNS disease 2 | NA | Median: 64,IQR 54-71 | Covid Death |
| Fernandes G et al.^28^ | Brazil | Cross sectional | April-August 2020 | 2021 | NA | 26 | 385 (Lung 18,Breast 93,Hematological 47 ) | NA | Female 234,Male 177 | NA | NA | NA | <60: 215,>60: 196 | Covid Death |
| Glasbey J et al.^29^ | International | Multi center,prospective cohort | April-June 2020 | 2020 | NA | 25 | 263 (Lung 25,Breast 24,Other 214) | 8683 (Gyncecological 1057) | Female 119,Male 169 | Early 181,Advance 107 | Pre-existing respiratory condition 45,Obese 56 | Minor Surgery 36,Major Surgery 252 | <50: 30,50-59: 39,60-69: 87,70-79: 96,>80: 36 | Covid Infection |
| Grivas P et al.^30^ | CCC19-International | Multi center,prospective cohort | March-November 2020 | 2021 | NA | 322 | 4796 (Lung 409,Breast 967,Hematological 1097) | NA | Female 2527,Male 2436 | NA | Cardiovascular 1582,Pulmonary 1091,Renal disease 831,Diabetes 1385 | Chemotherapy 802,Immunotherapy 248,Targeted therapy 693,Endocirne therapy 483,Locoregional therapy 422 | <65: 2282,65-74: 1309,>75: 1375 | Covid Hospitalization & Covid Death |
| Hathout L et al.^31^ | United States of America | Multi center,retrospective cohort | February-June 2020 | 2020 | NA | 3 (Cervical) | 0 | 44 (Endometrial 24, Cervical 12) | Female 3 | NA | Respiratory disease 1,Vascular disease 23,Respiratory+Vascular 4,HIV 3 | Brachytherapy 3 | NA | Covid Infection |
| Jee J et al.^32^ | United States of America | Single center,retrospective cohort | March-April 2020 | 2020 | NA | 15 (Cervical 2,Endometrial 6,Ovarian 5,Vaginal 1,Vulvar 1) | 294 (Lung 29 ,Breast 56,Hematological 71) | NA | Female 150,Male 159 | Metastasis 168 | Pulmonary disease 35,Cardiovascular disease 221,Metabolic disease 156,Neurologic disease 29,HIV 3,Liver disease 4 | Chemotherapy 102 | <60: 158,>60: 151 | Covid Death and Severe Covid |
| Johannesen T et al.^33^ | Norway | Multi center,retrospective cohort | January-May 2020 | 2021 | NA | 33 | 514 (Lung 13,Breast 85,Hematological 54) | 305299 (Gynecologic cancer 23827) | NA | Localized 36,Distant disease 6 | NA | SACT 71,Surgery 90,Radiotherapy 7 | NA | Covid Infection |
| Kulle C et al.^34^ | Turkey | Single center,retrospective cohort | March-June 2020 | 2021 | NA | 0 | 1 | 403 (Ovarian 14,Endometrial 9,Cervical 5,Uterine Sarcoma 1,Vulva 1) | NA | NA | NA | Surgery 1 | NA | Covid Infection |
| Kuru B et al.^35^ | Turkey | Single center,retrospective cohort | March-October 2020 | 2021 | 2 | 1 (Ovarian) | 0 | 61 | Female 3 | NA | Hypertension 2 | Surgery 1 | >65: 1,<65: 2 | Covid Infection |
| Kwon D et al. | United States of America | Multi center,retrospective cohort | February-December 2020 | pre-prints | NA | 119 | 1662 (Lung 33,Breast 241,Hematological 321) | 48137 (Gynecologic cancer 2877) | Female 950,Male 831 | NA | Heart disease 321,Pulmonary disease 294,CKD 273,Diabetes 474,Obese 481 | SACT 601,Hormonal therapy 86 | 18-65: 1044,65-75: 420,>75: 317 | Covid Infection |
| Lara O et al.^36^ | United States of America | Multi center,prospective cohort | March-June 2020 | 2021 | NA | 193 (Uterine 87,Epithelial Ovarian 62,Cervical 24,Vulva 8,Non-Epithelial Ovarian 3,Vaginal 3) | NA | NA | Female 193 | I/II: 74, III/IV: 100 | Hypertension 115,Diabetes 70,Asthma 21,COPD 5,Coronary artery disease 13,Autoimune disease 18,CKD 21 | Surgery 12,Radiotherapy 8,SACT 98 | Median 65,IQR 54-73 | Covid Hospitalization , Severe Covid & Covid Death |
| Lee L et al.^37^ | United Kingdom | Multi center,prospective cohort | March-April 2020 | 2020 | NA | 45 | 755 (Lung 90,Breast 102,Hematological 169) | NA | Female 349,Male 449 | Localized 149,Metastatic 347,Advanced stage 78 | Cardiovascular disease 109,COPD 61,Diabetes 131,Hypertension 247 | SACT 461,Surgery 29,Radiotherapy 76 | Median 69,IQR 59-76 | Covid Death |
| Lei S et al.^38^ | China | Multi center,retrospective cohort | January-February 2020 | 2020 | 25 | 1 (Ovarian) | 8 | NA | Female 20,Male 14 | NA | Hypertension 13,Diabetes 8,Cardiovascular disease 7,Cerebrovascular disease 2,COPD 1,CKD 1 | Surgery 9 | Median 55,IQR 43-63 | Covid Death |
| Li H et al.^39^ | United Kingdom | Multi center,retrospective cohort | March-October 2020 | 2021 | 275 | 17 (Uterine 7,Ovarian 10) | 272 (Lung 18,Breast 42,Hematological 53 ) | 4161 (Uterine 107,Ovarian 115 ) | Female 120,Male 168 | Localized 235,Metastasis 53 | NA | NA | 50-59: 28,60-69: 62,70-79: 159,80-84: 39 | Covid Infection & Covid Death |
| Liang J et al.^40^ | China | Single center,retrospective cohort | January-April 2020 | 2020 | NA | 10 (Uterine 4,Cervical 5,Ovarian 1) | 99 (Lung 14,Breast 11,Hematological 12) | NA | Female 52,Male 57 | I/II/III: 86, IV: 23 | Hypertension 38,Diabetes 18,Cardiovascular disease 10,Cerebrovascular disease 4,Chronic pulmonary disease 19,CKD 3,Chronic liver disease 10 | Surgery 69,Adjuvant 79,Chemo-radiation 71,Targeted-immunotherapy 12 | >65: 55,<65: 54 | Covid Death |
| Liu C et al.^41^ | China | Multi center,prospective cohort | December 2019 -March 2020 | 2020 | NA | 17 | 199 (Lung 49,Breast 34) | NA | Female 103,Male 113 | I-II: 83,III-IV: 85 | Diabetes 33,Hypertension 74,Cardiovascular 27,Cerebrovascular 18,COPD 21,Chronic liver disease 13,CKD 9 | 78 | Median 63,IQR 57-70,2 | Covid Death |
| Mehta V et al.^42^ | United States of America | Single center,retrospective cohort | March-April 2020 | 2020 | 1090 | 12 | 206 (Lung 11,Breast 29,Hematological 108) | NA | Female 91,Male 127 | Metastasis 42, Active cancer 92 | DM 80,Hypertension 147,Chronic lung disease 62,CKD 53,Coronary artery disease 43,CHF 33 | Chemotherapy 42,Immunotherapy 5,Radiotherapy 49 | 0-17: 3,18-44: 13,45-64: 64,65-74: 59,>75: 79 | Covid Death |
| Modi C et al.^43^ | United States of America | Multi center,prospective cohort | April-July 2020 | 2021 | NA | 1 | 4 (Lung 1,Breast 1) | 331 (Gynecologic 26) | Female 3,Male 2 | I-II: 3,III-IV: 2 | Comorbidity score^#^: 2: 2,5: 2,8: 1 | Radiotherapy 5 | <65: 3,>65: 2 | Covid Infection |
| Monroy-Iglesias MJ et al.^44^ | Italy | Multi center,prospective cohort | March-September 2020 | 2021 | NA | 2 | 14 | 3014 (Gynecological 382) | Female 2 | NA | NA | Surgery 16 | NA | Covid Infection, Severe Covid & Covid Death |
| Mousavi S et al.^45^ | Iran | Single center,retrospective cohort | February-April 2020 | 2021 | NA | 3 (Ovarian) | 30 (Lung 4,Breast 6) | NA | Female 15,Male 18 | I/II/III: 17,IV:16 | Cardiovascular & cerebrovascular disease 9,Diabetes 8,Chronic pulmonary disease 5,Chronic liver disease 1 | Cytotoxic chemotherapy 18 | Mean 63.9 | Covid Death |
| Nakamura S et al.^46^ | Japan | Single center,restrospective cohort | January-May 2020 | 2020 | NA | 1 (Cervical) | 31 (Lung 2,Breast 2,Hematological 7) | NA | Female10,Male 22 | Active cancer 17 | Diabetes 7,Hypertension 13,Coronary heart disease 4,COPD 4,Asthma 2 | Surgery 13,SACT 17 | >70: 20,<70: 12 | Covid Death |
| Ning M et al.^47^ | United States of America | Single center,prospective cohort | March-April 2020 | 2020 | NA | 2 (Endometrial 1,Vaginal 1 ) | 5 (Breast 1) | 114 (Gynecological 12) | Female 2 | Metastasis 2,III-IV: 2,Recurrent disease 2 | NA | Radiotherapy 7 | <65: 4,>65: 3 | death Covid, Covid Infection & Covid Death |
| OnCovid Study Group^48^ | OnCovid-Europe | Multi center,prospective cohort | February 2020-February 2021 | 2021 | NA | 115 | 2413 (Lung 11,Breast 29,Hematological 108 ) | NA | Female 1240,Male 1390 | Localized 1237,Advanced 1244 | 0-1: 1414 ,>2: 1220 | 1305 | <65: 1083, >65: 1538 | Covid Death |
| Ramaswamy A et al.^49^ | India | Single center,prospective cohort | April-June 2020 | 2020 | NA | 13 | 217 (Lung 12,Breast 30,Hematological 90) | NA | Female 106,Male 124 | Advanced 52,I-III: 93 | Diabetes 30,Hypertension 25,Cardiac illness 2 | SACT 230 | Median 42,IQR1-75 | Covid Death |
| Roel E et al.^50^ | Spain | Multi center,retrospective cohort | March-May 2020 | 2021 | 93558 | 436 (Corpus Uterus 291,Cervix 81,Ovary 64) | 4957 (Lung 159,Breast 1236,Hematological 513 ) | 255274 (Corpus Uterus 12665,Cervix 3232,Ovary 3564) | Female 57507,Male 41444 | NA | Autoimmune 6322,CKD 4167,COPD 2476,Heart disease 11076,Diabetes 6239,Obese 27840,Dementia 2011,Hyperlipidemia 11015 | NA | 18-39: 30648,40-59: 44909,60-69: 10602,70-79: 6419,>80: 6373 | Covid Infection, Covid Hospitalization & Covid Death |
| Russell B et al.^51^ | United Kingdom | Single center,retrospective cohort | March-June 2020 | 2021 | NA | 10 | 180 (Lung 22,Breast 27,Hematological 33) | 1962 | Female 78,Male 112 | I-II: 55,III-IV: 110 | NA | SACT 92,Combination therapy 11 | <50: 30,50-59: 36,60-69: 55,70-79: 40,>80: 29 | Covid Infection |
| Shi Z et al. | United Kingdom | Multi center,prospective cohort | June 2020 | pre-prints | 1306 | 9 (Cervix 2,Corpus Uteri 2,Ovary 5 ) | 409 (Lung 10,Breast 47,Hematological 49) | 2139 (Vulva 6,Cervix 7,Corpus Uteri 26,Ovary 20 ) | Female 746,Male 816 | NA | COPD 239,Asthma 240,Heart disease 672,Stroke 67,Hypertension 689,Obese 124,Diabetes 232 | NA | Cancer: mean 61.36,IQR 56.5-67.5, Non cancer: mean 56.11,IQR 47.5-64.5 | Covid Infection & Covid Death |
| Song C et al.^52^ | China | Multi center,retrospective cohort | December 2019 -March 2020 | 2020 | NA | 17 (Ovarian 3,Endometrial 4,Cervical 10) | 206 (Lung 39,Breast 31,Hematological 15) | NA | Female 107,Male 116 | NA | BMI >25: 30 | 126 | Median 63,IQR 56-71 | Covid Death |
| Song K et al.^53^ | China | Multi center,retrospective cohort | January-July 2020 | 2020 | NA | 10 | 238 (Lung 61,Breast 37) | NA | Female 120,Male 128 | I-III: 148,IV: 66 | Diabetes 38,Hypertension 83,Cardiovascular 28,Cerebrovascular 18,COPD 21 | Surgery 25,Radiotherapy 10,Combined 15,SACT 51 | Median 63,IQR 57-70 | Covid Death and Severe Covid |
| Tian J et al.^54^ | China | Multi center,retrospective cohort | January-March 2020 | 2020 | 519 | 15 (Cervical 11,Endometrial 3,Ovarian 1) | 217 (Lung 23,Breast 31,Hematological 12) | NA | Female 379,Male 372 | I-III: 192,IV: 34 | Hypertension 292,Diabtes 198,Coronary heart disease 74,CKD 23,Cerebrovascular disease 23,Hepatitis 10,COPD 4 | Surgery 197,Chemo/Radiotherapy 214,Targeted/Immunotherapy 32 | Median 64,IQR 57-69 | Covid Death and Severe Covid |
| Villegas A et al.^55^ | Spain | Single center,retrospective cohort | March-April 2020 | 2020 | NA | 1 (Ovarian) | 6 | 138 | Female 2,Male 1 | Advance 1,Initial staging 1,Recurrence 1 | NA | NA | >65: 3 | Covid Infection & Covid Death |
| Wang Q et al.^56^ | United States of America | Multi center,Case control | August 2020 | 2020 | NA | 30 (Endometrial) | 1440 (Lung 140,Breast 370,Hematological 220) | 3070260 (Endometrial 41710) | Female 9700,Male 6830 | NA | NA | NA | <18: 20,18-65: 11610,>65: 3900 | Covid Infection |
| Yang F et al.^57^ | China | Single center,retrospective cohort | January-April 2020 | 2020 | NA | 6 (Cervical 4,Endometrial 1,Ovarian 1 ) | 46 (Lung 10,Breast 9 ) | NA | Female 24,Male 28 | NA | Hypertension 17,Diabetes 7,Coronary heart disease 5,Cerebrovascular disease 4,COPD 4,CKD 1,Cirrhosis 1 | Chemotherapy 6, Surgery 2,Immunotherapy 1 | <60: 20,>60: 32 | Covid Death |
| Yang K et al.^58^ | China | Multi center,retrospective cohort | January-March 2020 | 2020 | NA | 9 (Cervical ) | 142 (Lung 24,Breast 40,Hematological 22) | NA | Female 109,Male 96 | I-II: 109,III-IV: 40 | Hypertension 67,Diabetes 22,COPD 5,Coronary heart disease 16,CKD 4 | Surgery 140,Radiotherapy 37,SACT 129 | <60: 86,>60:119 | Covid Death |
| Yang S et al.^59^ | China | Single center,retrospective cohort | January 2020 | 2020 | 1 | 2 (Ovarian 1,Cervical 1) | NA | 31 | Female 3 | I: 1, III: 1 | Diabetes & Hypertension 2,Hypertension 1 | Surgery 2 | >45: 3 | Covid Infection |
| Zhang L et al.^60^ | China | Multi center,retrospective cohort | January-February 2020 | 2020 | NA | 3 (Ovary 1,Endometrial 1,Cervix 1 ) | 25 (Lung 7,Breast 3 ) | NA | Female 11,Male 17 | I/II/III: 18,IV: 10 | Diabetes 4,Cardio&Cerebrovascular disease 4,Chronic pulmonary disease 1,Chronic liver disease 2 | Surgery 21,Chemo/radiotherapy 25,Target/Immunotherapy 6 | Median 65,IQR 56-70 | Covid Death and Severe Covid |
| Zhou K et al.^61^ | France | Multi center,retrospective cohort | June-November 2020 | 2021 | NA | 5 | 65 (Lung 8,Breast 36) | 808 (Gynecological 81) | Female 56,Male 14 | Localized 19,Locally advanced 9,Metastasis 32 | Hypertension 18,Diabetes 6,CKD 7,Heart failure 2,Autoimmune disease 2 | SACT 70,Radiotherapy 2,Surgery 4 | Median 61,IQR 27-81 | Covid Infection |

*Covid-19 population ; ^#^Charlson comorbidity index

CKD(Chronic Kidney Disease) ; COPD(Chronic Obstructive Pulmonary Disease) ; IQR(Interquartile Range) ; NA(Not Addressed) ; SACT(Systemic Anti-Cancer Therapy)
